# Global Safety Assessment of Adverse Events of Special Interest Following 2 Years of Use and 772 Million Administered Doses of mRNA-1273

**DOI:** 10.1101/2023.07.13.23291675

**Authors:** Veronica Urdaneta, Daina B. Esposito, Priyadarshani Dharia, Margot Stam Moraga, Kate Anteyi, Titi Oduyebo-Omotosho, Melissa Rossi, Paul Burton, José M. Vega, Rachel Dawson, Walter Straus

## Abstract

**Background:** The large-scale use of mRNA COVID-19 vaccines during the pandemic was associated with enhanced safety monitoring to ensure accurate and timely review of safety. We reviewed the mRNA-1273 (original strain) safety profile following 2 years of use (>772 million administered doses), primarily focusing upon predefined safety topics (ie, adverse events of special interest [AESI]) proposed in advance of COVID-19 vaccine use.

**Methods:** Cumulative mRNA-1273 safety data from spontaneous adverse event (AE) cases reported to the Moderna, Inc., global safety database (GSDB) between December 18, 2020, and December 17, 2022, were included. Reporting rates of AESI were calculated per 1 million doses of mRNA-1273 administered. Observed-to-expected (OE) ratios were computed by comparing observed rates of AESI to the background/expected rate for these events to evaluate potential associations with mRNA-1273.

**Results:** There were 658,767 identified case reports, associated with 2,517,669 AEs. Most AEs were non-serious (83.4%); 0.7% were fatal. AESI represented 13.7% of all AEs, with reporting rates for most AESI below the expected background incidence. Exceptions included anaphylaxis (OE ratio 3 days after vaccination, 2.19 [95% CI, 2.02-2.37]), myocarditis (OE ratio 7 days after vaccination, 1.41 [1.32-1.51]; among men aged 12-40 years, 9.75 [7.74-12.3]; and individuals aged 12-40 years, 3.51 [3.19-3.86]), and pericarditis (OE ratio 7 days after vaccination in individuals aged 12-40 years, 2.54 [2.16-2.99]).

**Conclusions:** With the exceptions of anaphylaxis, myocarditis, and pericarditis, this safety analysis of mRNA-1273 did not find evidence to suggest an increased risk for AESI identified for enhanced monitoring ahead of COVID-19 vaccine use.

## Introduction

The introduction and authorization of COVID-19 vaccines has had a major impact on decreasing COVID-19–associated hospitalizations and deaths during the pandemic [1–4]. Among these were 2 first-in-class mRNA vaccines, mRNA-1273 (SPIKEVAX; Moderna, Inc., Cambridge, USA) and BNT162b2 (COMIRNATY; Pfizer Inc., New York, USA; BioNTech Manufacturing GmbH, Mainz, Germany). In a phase 3 randomized controlled trial of mRNA-1273, 2 doses were well-tolerated and highly effective against symptomatic infection [5]. Real-world studies of mRNA-1273 subsequently demonstrated vaccine effectiveness against the ancestral SARS-CoV-2 strain and emergent variants of concern in the general population and among those at high risk of severe disease [6–11]. From initial authorization through December 2022, >248 and >152 million doses have been administered in the United States and European Union, respectively [12].

Ensuring vaccine safety is essential to preserving public trust in vaccines, particularly during a pandemic while using a new vaccine platform [13, 14]. Newly introduced or expanded safety systems (eg, Biologics Effectiveness and Safety [BEST], Vaccine Safety [V-safe], and Vaccine monitoring Collaboration for Europe [VAC4EU]) have been implemented in the United States and the European Union to complement existing public health infrastructure and support reporting requirements for vaccine manufacturers [15, 16]. These post-authorization safety monitoring measures are designed to detect emergent and rare vaccine-related safety concerns not identified in clinical trials due to inclusion/exclusion criteria and limitations in sample size [13, 15, 17]. In May 2020, in anticipation of COVID-19 vaccine development, the Brighton Collaboration (in support of the Coalition for Epidemic Preparedness Innovations Safety Platform for Emergency vACcines [SPEAC]) introduced a list of relevant adverse events of special interest (AESI) to help guide clinical trial and post-marketing data analysis. These AESI were based on immune-mediated COVID-19 sequelae or events common to many licensed vaccines with the potential to be observed in recipients of COVID-19 vaccines [18].

Following emergency use authorization (EUA) of COVID-19 vaccines, data emerged suggesting an increased risk for several AESI after vaccination [19–21]. In December, 2020, reports emerged of allergic reactions (anaphylaxis) following receipt of the mRNA COVID-19 vaccines [22]. In April 2021, there were reports of myocarditis and/or pericarditis (inflammation of the myocardium and pericardium, respectively) in individuals who had received one of the authorized mRNA COVID-19 vaccines [19]. Also, at that time, the Advisory Committee for Immunization Practice (ACIP) presented data on cases of thrombosis with thrombocytopenia syndrome (TTS), a condition characterized by the formation of blood clots combined with low platelet count [23]. Several months later, the ACIP reviewed cases of Guillain-Barré syndrome (GBS; an autoimmune reaction primarily affecting peripheral nerves) following COVID-19 vaccination [24].

Herein, we review 2 years of safety data for mRNA-1273 from spontaneous AE case reports submitted to the Moderna, Inc., global safety database (GSDB) according to previously defined AESI, by which time >772 million doses of mRNA-1273 had been administered. This analysis also includes estimation of reporting rates and observed-to-expected (OE) analysis, a quantitative pharmacovigilance approach used to estimate whether observed rates of certain AESI after vaccination exceeded background rates [25].

## Methods

### Data sources

The Moderna GSDB collects safety data submitted by regulatory authorities, healthcare providers, and consumers where mRNA-1273 is authorized for use. Safety data obtained through sponsored clinical or observational studies and cases extracted from secondary sources (such as events identified in the literature [MEDLINE, EMBASE]) are also added to the GSDB. The Medical Dictionary for Regulatory Activities (MedDRA) comprising internationally standardized terminology was used to code reported events. As a noninterventional, retrospective database study using mostly AE reports submitted voluntarily to the Moderna GSDB, as part of routine post-authorization safety surveillance efforts required by regulatory authorities, a central institutional review board (IRB; Advarra) confirmed this study met criteria for an exemption from IRB oversight under 45 CFR 46.104(d)(4).

From December 18, 2020 (date of first international EUA issuance), to December 17, 2022, the GSDB was reviewed for valid case reports (ie, contains an identifiable reporter, identifiable patient, adverse reaction, and suspected product) according to the list of AESI (published May 2020 and updated October 2022) [26], using standard and/or customized queries from the most updated version of the MedDRA (v25.1) and excluding clinical trial data. Reported AEs following immunization may include any untoward medical occurrence that follows administration and that does not necessarily have a causal relationship with vaccination [27]. This analysis was restricted to reports received for the mRNA-1273 (original strain) vaccine.

### Assessment of case reports

AESI were continuously and systematically reviewed by Moderna, Inc., as part of the standard process of identification and evaluation of possible safety risks associated with mRNA-1273. Formal pharmacovigilance case definitions were used to categorize reported AEs/AESI. A serious AE was defined as one that results in death, hospitalization, disability/permanent damage, or other event(s) jeopardizing the patient that might require medical/surgical intervention [28].

### Statistical analysis

AEs reported to the GSDB were reviewed and used for this analysis. Reported cases of AESI were characterized by age, sex, MedDRA preferred term (PT), time to onset (TTO) after vaccination, and dose number using descriptive statistics. Reporting rates were used to estimate the frequency of specific AESI, calculated as number of reported cases per 1 million doses administered. In this analysis, the doses administered were conservatively estimated as 55% of doses distributed globally. Vaccine exposure in different age groups and by sex was estimated based on the published distributions of COVID-19 vaccine recipient demographics in the United States published by the Centers for Disease Control as of December 17, 2022 [29].

### OE analysis

OE ratios were determined for each of the AESI included in this analysis. Expected incidence rates were identified from published sources [30–44], with an emphasis on recent, high-quality, large, population-based studies. For estimation of observed reporting rates, all reports were included regardless of causality or TTO. An assumed risk window of 21 days was assigned after each administered vaccine dose to estimate exposed person-time at risk, unless otherwise specified. This window was selected for consistency with analyses that have been conducted by the US Vaccine Safety Datalink. The sum of all person-time was then used as a denominator to calculate a reporting rate comparable to expected rates from published sources. These expected rates were then multiplied by the estimated exposed person-time to identify the count of expected cases [25]. The observed reporting rate for each AESI was divided by the expected rate and presented alongside its associated 95% confidence interval (CI), calculated as: e^(log(IRR)±1.96*SE(log(IRR)))^ [25]. Age- and sex-stratified assessments of OE ratios were also performed. Subpopulation evaluations presented in this analysis included children (<18 years), adults (18-64 years), and elderly individuals (_≥_65 years). For anaphylaxis, a 3-day risk window was applied to person-time and case counts; a 7-day risk window, stratified by dose, was used for myocarditis and pericarditis.

For any AESI where the lower bound of the 95% CI was >1 in either overall or subgroup analyses, cases were medically reviewed to determine the plausibility of an association. In some instances, expected background incidence rates could not reasonably be determined because of sparse or absent epidemiologic data in the general population, resulting in more qualitative review of case characteristics.

This OE analysis reports on all AESI initially proposed by SPEAC for COVID-19 vaccines and provides additional information on those that received attention by the ACIP during the first 6 months following emergency use: anaphylaxis, myocarditis, pericarditis, TTS, and GBS.

## Results

### Cases identified

As of December 17, 2022, 1,315,589,716 doses of mRNA-1273 have been distributed to 91 countries, with ∼772,908,958 doses administered. During this period, 658,767 cases were captured in the mRNA-1273 GSDB, which included 2,517,669 AEs (AESI represented 13.7% of AEs; **Table 1**). The overall reporting rate was 852.2 cases per 1 million doses administered.

**Table 1.**
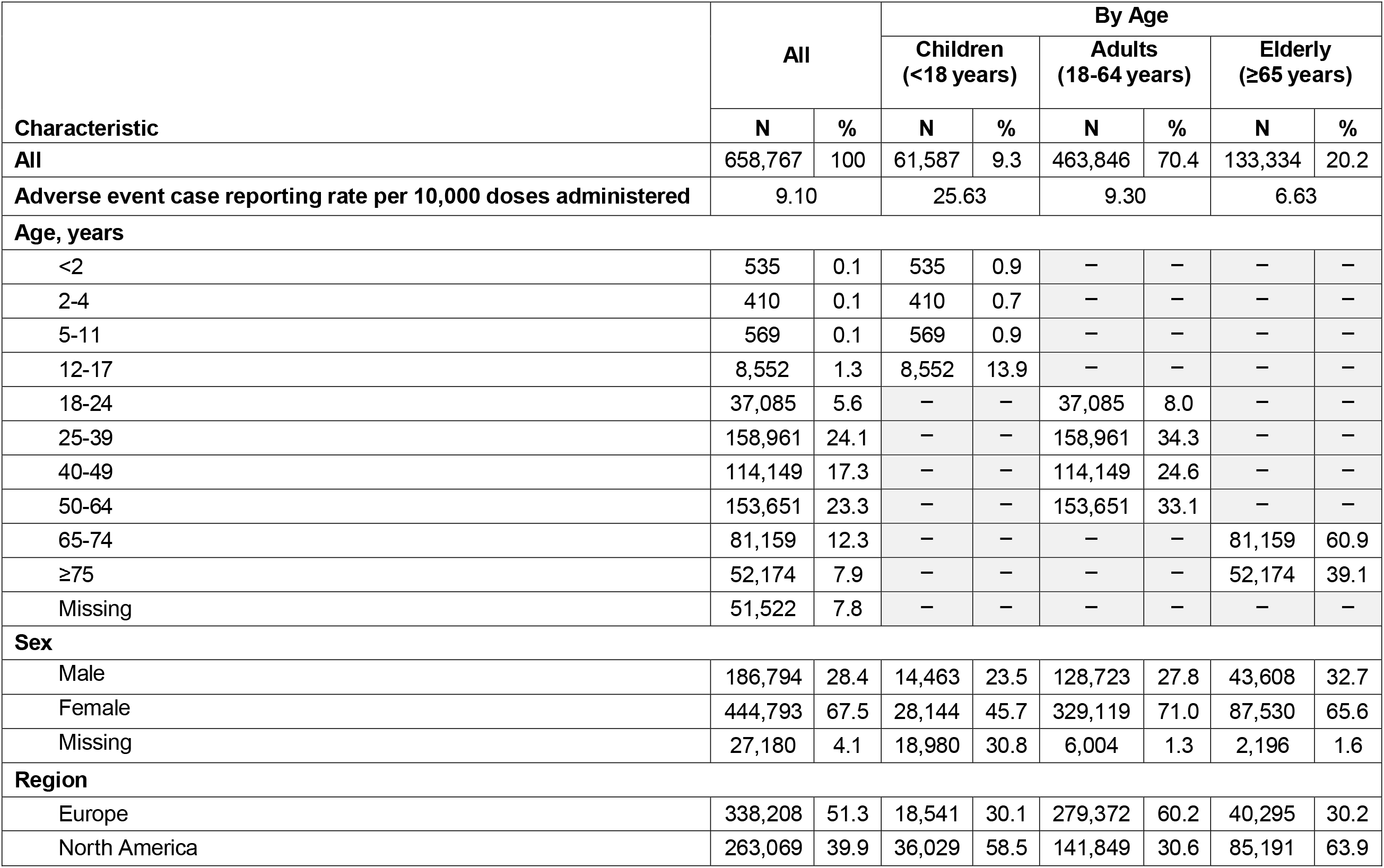
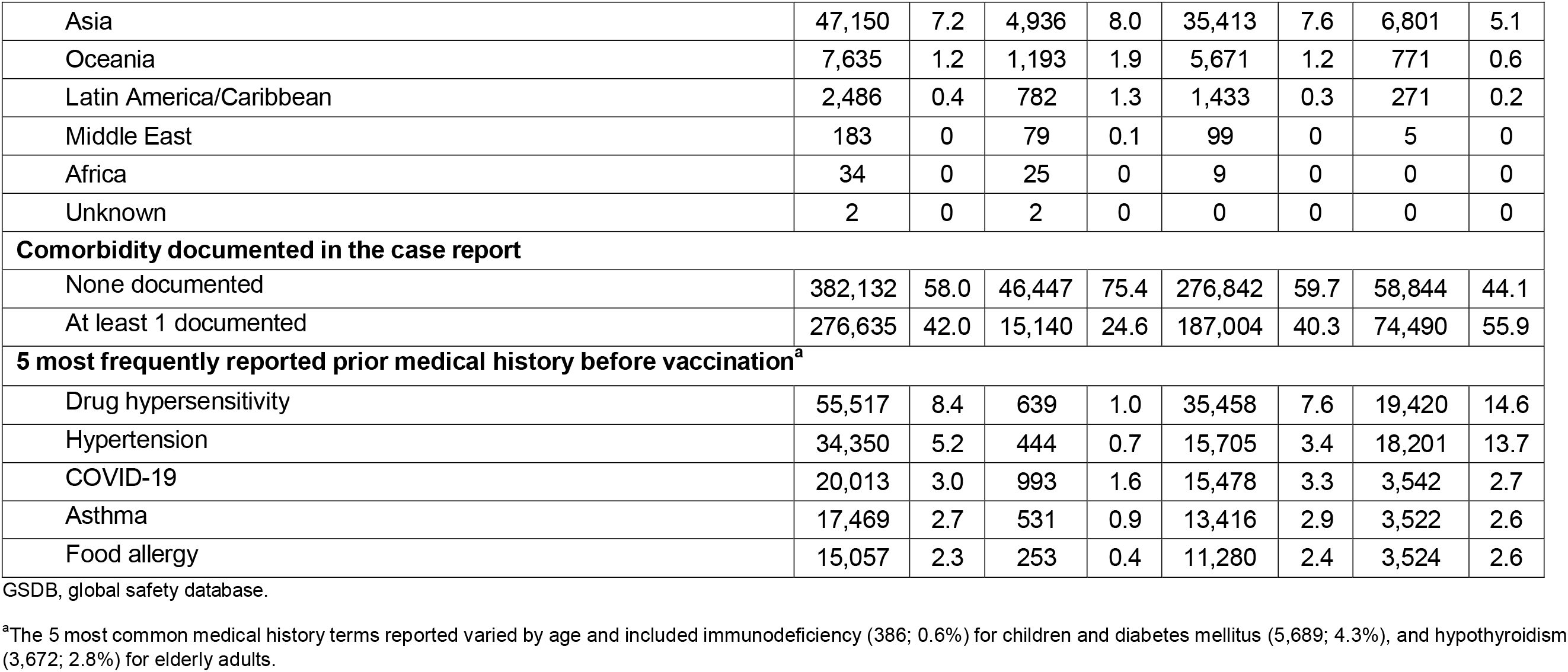
Characteristics of Cases Reported to the Moderna GSDB From December 2020 to December 2022.

In the first 6 months of vaccine distribution, the number of reported events increased as the number of doses distributed increased, with peak reported events reaching 344,842 in May 2021 (**Figure 1**). Since then, reported events gradually declined, with 16,706 events in December 2022.

**Figure 1.**
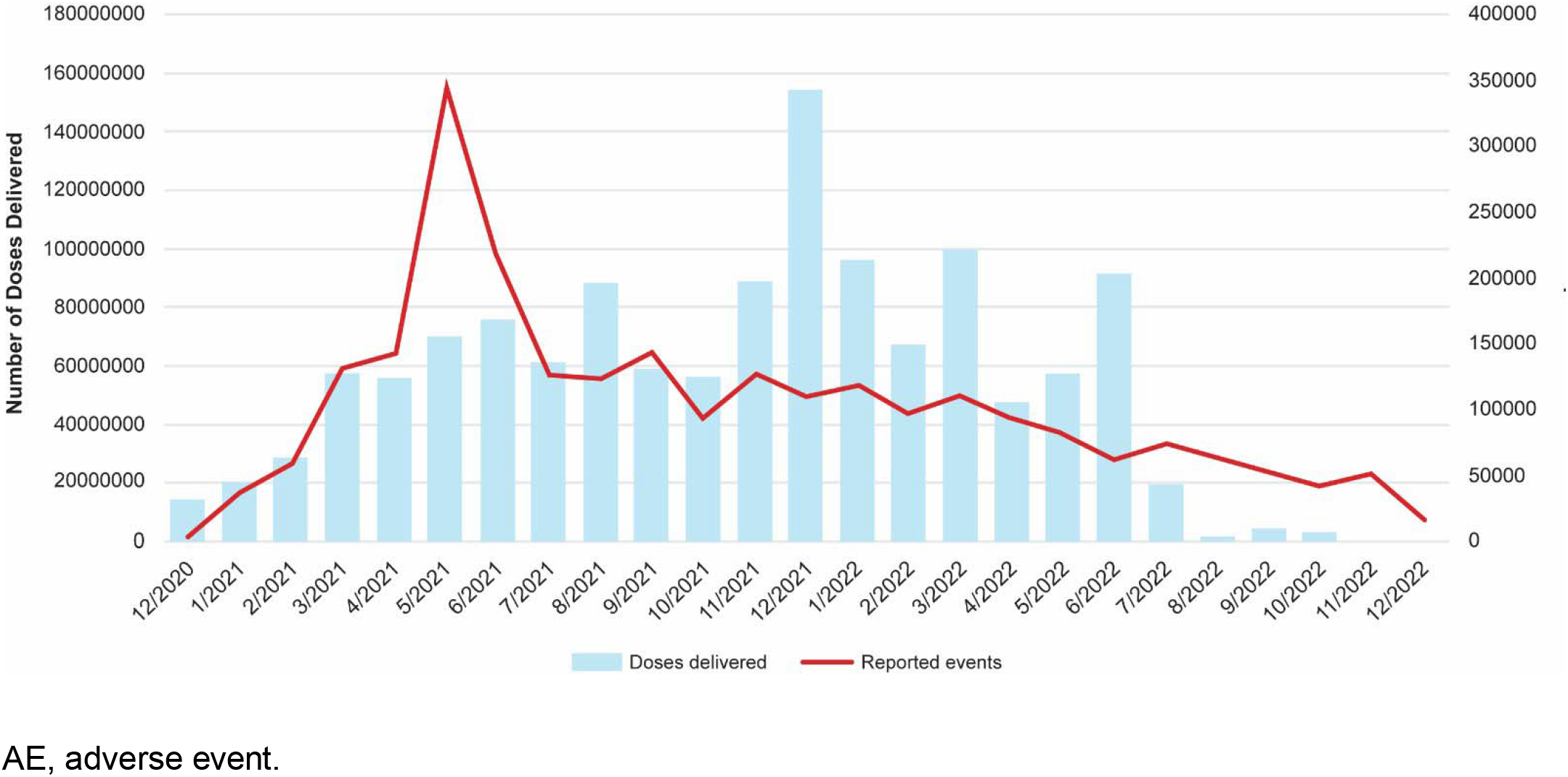
mRNA-1273 Doses Distributed and Reported Adverse Events by Month From December 2020 to December 2022. AE, adverse event.

Most cases were reported in individuals aged 18-64 years (70.4%), from Europe (51.3%) and North America (39.9%), and in women (67.5%). In 42% of cases, _≥_1 underlying health condition was present before vaccination; the most common conditions across age groups were drug hypersensitivity (8.4%), hypertension (5.2%), COVID-19 (3.0%), asthma (2.7%), and food allergy (2.3%; **Table 1**).

Most reported AEs were non-serious (83.4%) and reported by regulatory health authorities (79.7%; **Table 2**). The most frequently reported AEs in the overall population are summarized in **Table 2**. Of the reported AEs, 49.0% were associated with reactogenicity. More AEs were reported after dose 1 (30.1%) than subsequent doses. The median TTO from the most recent dose was 3 days (**Figure 2**). At the time of receipt, 47.4% of AEs had reported outcomes as recovered or recovering. Less than 1% of all AEs (0.7%) were fatal; this varied by age, with 70.4% of fatal AEs occurring in elderly adults. Cause of death was not reported for 16.9% of all fatal events. For those cases providing cause of death the leading event was COVID-19 (3.9% of all fatal events), followed by dyspnea (3.0% of all fatal events) and cardiac arrest (2.6% of all fatal events).

**Figure 2.**
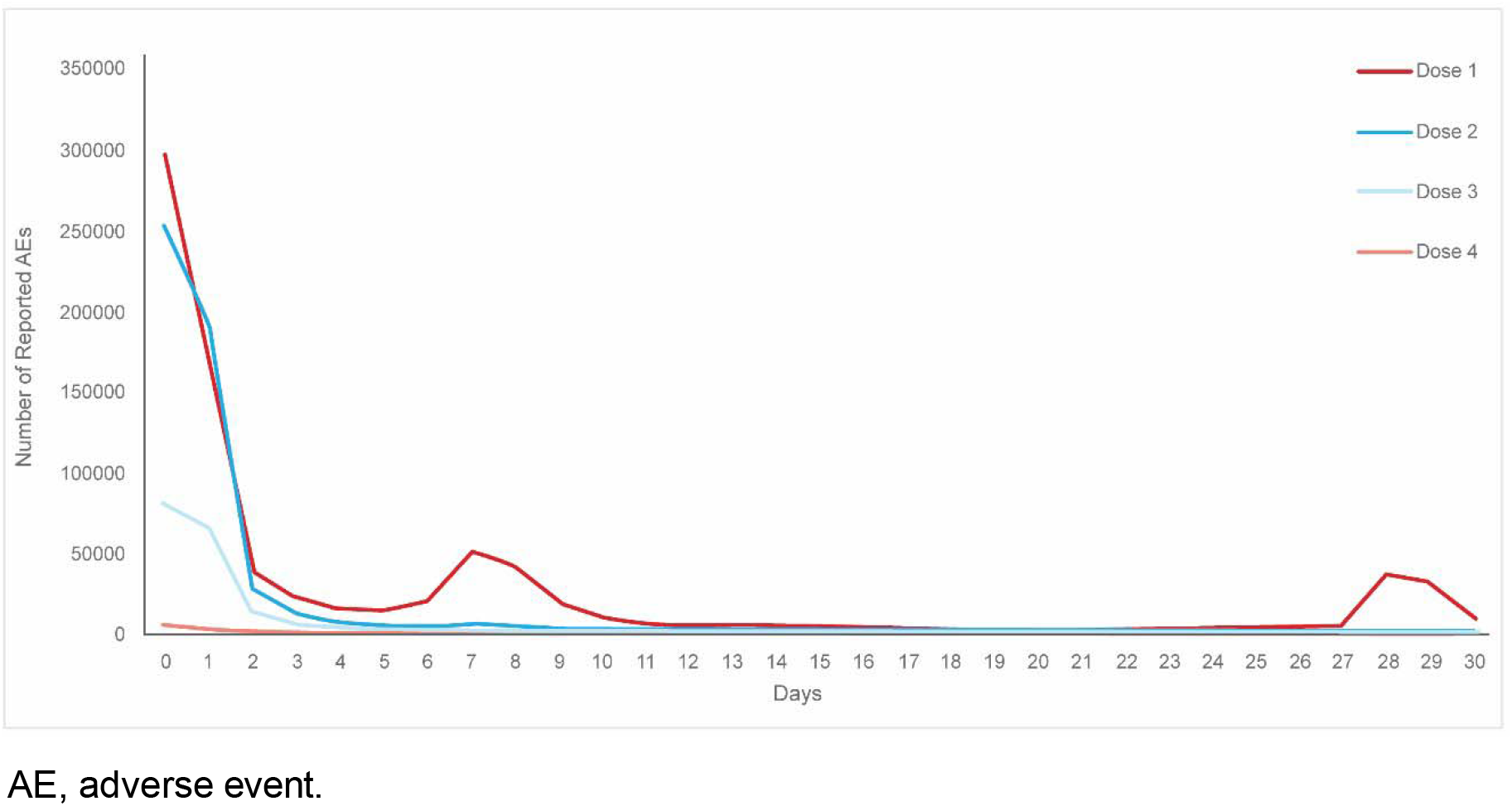
Time to Onset of Reported Adverse Events With mRNA-1273 by Dose. AE, adverse event.

**Table 2.** Characteristics of Adverse Events Reported to the Moderna GSDB From December 2020 to December 2022.

| Characteristic | All |  | By Age |  |  |  |  |  |
| --- | --- | --- | --- | --- | --- | --- | --- | --- |
|  |  |  | Children (<18 years) |  | Adults (18-64 years) |  | Elderly (≥65 years) |  |
|  | N | % | N | % | N | % | N | % |
| **All** | 2,517,669 | 100 | 150,768 | 6.0 | 1,890,309 | 75.1 | 476,592 | 18.9 |
| **Event seriousness<sup>a</sup>** |  |  |  |  |  |  |  |  |
| Serious | 418,715 | 16.6 | 25,830 | 17.1 | 291,077 | 15.4 | 101,808 | 21.4 |
| Non-serious | 2,098,954 | 83.4 | 124,938 | 82.9 | 1,599,232 | 84.6 | 374,784 | 78.6 |
| **Healthcare provider submitted the report** | 1,074,760 | 42.7 | 50,130 | 33.2 | 755,523 | 40.0 | 269,107 | 56.5 |
| **Reporter type** |  |  |  |  |  |  |  |  |
| Spontaneous report received from regulatory authority | 2,007,048 | 79.7 | 70,279 | 46.6 | 1,623,913 | 85.9 | 312,856 | 16.6 |
| Spontaneous report received directly from HCPs and consumers | 502,516 | 20.0 | 79,063 | 52.4 | 260,874 | 13.8 | 162,579 | 8.6 |
| Other (eg, literature) | 8,105 | 0.3 | 1,426 | 0.9 | 5,522 | 0.3 | 1,157 | 0.1 |
| **Dose number** |  |  |  |  |  |  |  |  |
| Dose 1 | 758,268 | 30.1 | 31,905 | 21.2 | 563,405 | 29.8 | 162,958 | 34.2 |
| Dose 2 | 606,530 | 24.1 | 14,712 | 9.8 | 470,410 | 24.9 | 121,408 | 25.5 |
| Dose 3 | 202,370 | 8.0 | 9,969 | 6.6 | 164,262 | 8.7 | 28,139 | 5.9 |
| Dose 4 | 11,467 | 0.5 | 704 | 0.5 | 4,251 | 0.2 | 6,512 | 1.4 |
| Other/not reported | 939,034 | 37.3 | 93,478 | 62.0 | 687,981 | 36.4 | 157,575 | 33.1 |
| **Event outcome at the time of report<sup>b</sup>** |  |  |  |  |  |  |  |  |
| Fatal | 17,751 | 0.7 | 527 | 0.3 | 4,723 | 0.2 | 12,501 | 2.6 |
| Recovered/recovering | 1,192,627 | 47.4 | 57,107 | 37.9 | 944,247 | 50.0 | 191,273 | 40.1 |
| Not recovered | 709,987 | 28.2 | 27,227 | 18.1 | 562,760 | 29.8 | 120,000 | 25.2 |
| Not reported | 597,304 | 23.7 | 65,907 | 43.7 | 378,579 | 20.0 | 152,818 | 32.1 |
| **Most frequently reported events** |  |  |  |  |  |  |  |  |
| Headache | 146,039 | 5.8 | 7,197 | 4.8 | 119,374 | 6.3 | 19,468 | 4.1 |
| Pyrexia | 139,460 | 5.5 | 8,405 | 5.6 | 111,181 | 5.9 | 19,874 | 4.2 |
| Fatigue | 127,184 | 5.1 | 5,549 | 3.7 | 102,437 | 5.4 | 19,198 | 4.0 |
| Chills | 97,683 | 3.9 | 4,142 | 2.8 | 77,877 | 4.1 | 15,664 | 3.3 |
| Myalgia | 91,510 | 3.6 | 4,339 | 2.9 | 75,322 | 4.0 | 11,849 | 2.5 |
| Injection site pain | 70,431 | 2.8 | 1,497 | 1.0 | 61,471 | 3.3 | 7,463 | 1.6 |
| Nausea | 69,913 | 2.8 | 3,233 | 2.1 | 55,846 | 3.0 | 10,834 | 2.3 |
| Malaise | 67,283 | 2.7 | 2,347 | 1.6 | 56,903 | 3.0 | 8,033 | 1.7 |
| Arthralgia | 55,998 | 2.2 | 2,381 | 1.6 | 44,327 | 2.3 | 9,290 | 2.0 |
| Pain in extremity | 53,606 | 2.1 | 3,782 | 2.5 | 37,563 | 2.0 | 12,261 | 2.6 |

AE, adverse event; GSDB, global safety database; HCP, healthcare provider.
<sup>a</sup>A serious AE was defined as one that results in death, hospitalization, disability/permanent damage, or other event(s) jeopardizing the patient that might require medical/surgical intervention.
<sup>b</sup>For spontaneous case reports, event outcomes were those provided at the time of submission of the adverse event report and often did not contain important elements of the adverse event, including any follow-up information.

### AESI

#### Anaphylaxis

Based on cumulative exposure and published background rates, ∼6,277 cases of anaphylaxis were expected <21 days post-vaccination. There were 2,588 reported cases (2,651 events) of anaphylaxis (0.4% of all reported cases); most were in women (73.6%). Of these cases, 67 (2.6%) reported a fatal outcome; however, most cases (98.5%) were unrelated to vaccination. When examined by dose number, 47.0% of cases occurred after dose 1, 19.7% after dose 2, 8.3% after dose 3, and 0.8% after dose 4; 24.2% did not indicate dose number. Most events occurred on the same day of vaccination (63.3%) regardless of the dose number.

The reporting rate of anaphylaxis was 3.58 cases per 1 million doses administered, with a higher rate in adults (18-64 years old; 4.23 cases per 1 million doses; **Table 3**). When considering a standard 21-day interval assessed across AESI, the reporting rate was below published population-based rates (OE ratio, 0.41; 95% CI, 0.39-0.43). However, when comparisons considered events within a 3-day risk window (more plausible time frame for this AE), there was an overall increase above background rates (OE ratio, 2.19; 95% CI, 2.02-2.37), among women (3.13; 95% CI, 2.82-3.47), and in adults (18-64 years of age; 4.01; 95% CI, 3.60-4.46). Consideration of background rates from alternate published studies produced consistent conclusions [43].

**Table 3.** Reporting Rates of AESI by Age and Sex From December 2020 to December 2022.

| AESI | Reporting Rate per 1 Million Doses Administered |  |  |  |  |  | % of Total Cases Reported | OE Ratio (95% CI)<sup>a</sup> |
| --- | --- | --- | --- | --- | --- | --- | --- | --- |
|  | All | Sex |  | Age (years) |  |  |  |  |
|  |  | Male | Female | <18 | 18 to 64 | ≥65 |  |  |
| Acute respiratory distress | 0.85 | 1.00 | 0.71 | 0.00 | 0.34 | 2.19 | 0.093 | 0.05 (0.05-0.05) |
| Multisystem inflammatory syndrome | 0.23 | 0.21 | 0.25 | 0.21 | 0.18 | 0.31 | 0.025 | 0.16 (0.13-0.18) |
| Myocarditis and pericarditis |  |  |  |  |  |  |  |  |
| Myocarditis with or without pericarditis (21-day risk window) | 6.07 | 9.41 | 2.81 | 8.07 | 6.85 | 1.27 | 0.666 | 0.93 (0.89-0.97) |
| Myocarditis with or without pericarditis (7-day risk window) | 3.07 | 5.20 | 1.15 | 5.49 | 3.95 | 0.44 | 0.003 | 1.41 (1.32-1.51) |
| Myocarditis with or without pericarditis (7-day risk window) in those aged 12-40 years | 8.49 | 13.37 | 2.03 | - | - | - | 0.002 | 3.51 (3.19-3.86) |
| Dose 1 (7-day risk window) in those aged 12-40 years | 0.55 | - | - | - | - | - | 0.000 | 1.13 (0.95-1.34) |
| Males | - | 0.89 | - | - | - | - | 0.000 | 1.82 (1.46-2.28) |
| Females | - | - | 0.23 | - | - | - | 0.000 | 0.47 (0.35-0.64) |
| Dose 2 (7-day risk window) in those aged 12-40 years | 1.14 | - | - | - | - | - | 0.001 | 5.24 (4.43-6.19) |
| Males | - | 2.12 | - | - | - | - | 0.001 | 9.75 (7.74-12.3) |
| Females | - | - | 0.23 | - | - | - | 0.000 | 1.04 (0.77-1.39) |
| Dose 3 (7-day risk window) in those aged 12-40 years | 0.22 | - | - | - | - | - | 0.000 | 1.49 (1.2-1.85) |
| Males | - | 0.38 | - | - | - | - | 0.000 | 2.49 (1.87-3.31) |
| Females | - | - | 0.09 | - | - | - | 0.000 | 0.57 (0.39-0.84) |
| Pericarditis without myocarditis (21-day risk window) | 4.72 | 6.18 | 3.29 | 3.08 | 3.25 | 0.81 | 0.003 | 0.79 (0.75-0.83) |
| Pericarditis without myocarditis (7-day risk window) | 1.33 | 1.57 | 1.11 | 0.96 | 1.66 | 0.44 | 0.001 | 1.02 (0.93-1.12) |
| Pericarditis without myocarditis (7-day risk window) in those aged 12-40 years | 2.27 | 2.88 | 1.70 | - | - | - | 0.001 | 2.54 (2.16-2.99) |
| Dose 1 (7-day risk window) in those aged 12-40 years | 0.23 | - | - | - | - | - | 0.000 | 1.28 (0.97-1.69) |
| Males | - | 0.28 | - | - | - | - | 0.000 | 1.54 (1.05-2.25) |
| Females | - | - | 0.19 | - | - | - | 0.000 | 1.05 (0.71-1.56) |
| Dose 2 (7-day risk window) in those aged 12-40 years | 0.22 | - | - | - | - | - | 0.000 | 2.71 (2.02-3.64) |
| Males | - | 0.31 | - | - | - | - | 0.000 | 3.88 (2.58-5.84) |
| Females | - | - | 0.13 | - | - | - | 0.000 | 1.6 (1.03-2.5) |
| Dose 3 (7-day risk window) in those aged 12-40 years | 0.06 | - | - | - | - | - | 0.000 | 1.08 (0.74-1.59) |
| Males | - | 0.09 | - | - | - | - | 0.000 | 1.53 (0.91-2.55) |
| Females | - | - | 0.04 | - | - | - | 0.000 | 0.68 (0.37-1.23) |
| Other forms of cardiac injury |  |  |  |  |  |  |  |  |
| Arrhythmia | 12.16 | 10.59 | 13.22 | 1.46 | 11.85 | 12.59 | 1.336 | 0.04 (0.03-0.04) |
| Heart failure | 1.90 | 2.22 | 1.58 | 0.00 | 1.07 | 4.04 | 0.209 | 0.02 (0.02-0.02) |
| Ischemic coronary artery disease | 2.49 | 3.29 | 1.70 | 0.04 | 1.87 | 3.88 | 0.274 | 0.01 (0.01-0.01) |
| Microangiopathy | 0.04 | 0.05 | 0.03 | 0.00 | 0.01 | 0.04 | 0.004 | 0.10 (0.07-0.14) |
| Stress cardiomyopathy | 0.08 | 0.03 | 0.13 | 0.00 | 0.04 | 0.18 | 0.009 | 0.03 (0.02-0.04) |
| Any thromboembolic event | 17.53 | 18.02 | 16.53 | 0.62 | 13.78 | 26.19 | 1.926 | 0.12 (0.12-0.12) |
| Hemorrhagic disorders including DIC |  |  |  |  |  |  |  |  |
| Deep vein thrombosis | 7.13 | 7.40 | 6.75 | 0.37 | 6.21 | 9.58 | 0.783 | 0.02 (0.02-0.02) |
| Disseminated intravascular coagulation | 0.06 | 0.04 | 0.07 | 0.00 | 0.05 | 0.07 | 0.006 | 0.01 (0.01-0.01) |
| Cerebral venous sinus thrombosis | 0.28 | 0.27 | 0.28 | 0.04 | 0.31 | 0.20 | 0.031 | 0.19 (0.17-0.23) |
| Pulmonary embolism | 4.37 | 4.49 | 4.18 | 0.17 | 3.68 | 6.19 | 0.480 | 0.04 (0.04-0.04) |
| Splanchnic venous thrombosis | 0.10 | 0.12 | 0.07 | 0.00 | 0.06 | 0.05 | 0.010 | 0.05 (0.04-0.06) |
| Stroke, all | 6.24 | 6.44 | 5.87 | 0.37 | 3.94 | 11.70 | 0.685 | 0.05 (0.05-0.05) |
| Stroke, hemorrhagic | 1.03 | 1.06 | 0.97 | 0.17 | 0.68 | 1.89 | 0.113 | 0.01 (0.01-0.01) |
| Anosmia/ageusia | 4.51 | 2.92 | 5.64 | 0.58 | 4.57 | 3.69 | 0.495 | 0.59 (0.56-0.61) |
| Chilblain-like lesions | 0.15 | 0.08 | 0.20 | 0.04 | 0.15 | 0.09 | 0.016 | 0.02 (0.02-0.02) |
| Erythema multiforme<sup>b</sup> | 0.47 | 0.37 | 0.52 | 0.67 | 0.53 | 0.21 | 0.051 | 0.10 (0.09-0.12) |
| Single organ cutaneous vasculitis | 0.51 | 0.33 | 0.64 | 0.12 | 0.47 | 0.58 | 0.056 | 0.16 (0.14-0.17) |
| Acute kidney injury | 1.49 | 1.69 | 1.29 | 0.17 | 0.75 | 3.31 | 0.163 | 0.01 (0.01-0.01) |
| Acute liver injury | 0.52 | 0.42 | 0.59 | 0.12 | 0.47 | 0.62 | 0.058 | 0.11 (0.10-0.12) |
| Acute pancreatitis | 0.38 | 0.38 | 0.37 | 0.00 | 0.35 | 0.45 | 0.042 | 0.14 (0.12-0.15) |
| Rhabdomyolysis | 0.24 | 0.25 | 0.21 | 0.00 | 0.18 | 0.36 | 0.026 | 0.11 (0.09-0.13) |
| Subacute thyroiditis<sup>c</sup> | 0.62 | 0.25 | 0.94 | 0.08 | 0.78 | 0.19 | 0.068 | 0.04 (0.03-0.04) |
| Anaphylaxis |  |  |  |  |  |  |  |  |
| Anaphylaxis (21-day risk window) | 3.58 | 1.83 | 4.97 | 0.92 | 4.23 | 1.67 | 0.393 | 0.41 (0.39-0.43) |
| Anaphylaxis (3-day risk window) | 2.71 | 1.31 | 3.88 | 0.67 | 3.38 | 1.05 | 0.003 | 2.19 (2.02-2.37) |
| Thrombocytopenia | 3.26 | 2.51 | 3.78 | 0.50 | 0.90 | 1.82 | 0.358 | 0.07 (0.07-0.07) |
| Generalized convulsion | 4.68 | 4.22 | 4.84 | 3.29 | 5.26 | 2.78 | 0.515 | 0.16 (0.16-0.17) |
| Acute disseminated encephalomyelitis | 0.15 | 0.15 | 0.14 | 0.12 | 0.16 | 0.11 | 0.016 | 0.41 (0.33-0.52) |
| Guillain-Barré syndrome | 0.94 | 1.03 | 0.83 | 0.21 | 0.86 | 1.09 | 0.104 | 0.26 (0.24-0.29) |
| Acute aseptic arthritis | 4.10 | 2.34 | 5.50 | 0.12 | 3.87 | 4.18 | 0.450 | 0.08 (0.08-0.08) |
| Aseptic meningitis | 0.22 | 0.20 | 0.23 | 0.04 | 0.24 | 0.14 | 0.024 | 0.04 (0.03-0.05) |
| Encephalitis/encephalomyelitis |  |  |  |  |  |  |  |  |
| Encephalitis | 0.39 | 0.38 | 0.38 | 0.17 | 0.38 | 0.37 | 0.042 | 0.35 (0.31-0.40) |
| Multiple sclerosis | 0.51 | 0.24 | 0.74 | 0.04 | 0.61 | 0.22 | 0.056 | 0.06 (0.05-0.06) |
| Myasthenia gravis | 0.24 | 0.29 | 0.18 | 0.04 | 0.13 | 0.46 | 0.026 | 0.48 (0.40-0.58) |
| Optic neuritis | 0.19 | 0.14 | 0.23 | 0.12 | 0.22 | 0.08 | 0.021 | 0.07 (0.06-0.08) |
| Fibromyalgia | 0.45 | 0.07 | 0.75 | 0.00 | 0.46 | 0.38 | 0.049 | 0.01 (0.01-0.01) |
| Transverse myelitis | 0.17 | 0.17 | 0.17 | 0.00 | 0.14 | 0.21 | 0.019 | 0.23 (0.19-0.28) |
| Bell's palsy | 4.67 | 4.16 | 4.99 | 0.79 | 4.83 | 4.13 | 0.513 | 0.10 (0.10-0.10) |
| Thrombocytopenia and thrombosis | 0.32 | 0.34 | 0.28 |  |  |  | 0.035 | 0.27 (0.24-0.31) |
| Immune thrombocytopenia | 0.49 | 0.40 | 0.56 | 0.33 | 0.43 | 0.59 | 0.054 | 0.11 (0.10-0.12) |
| Capillary leak syndrome<sup>d</sup> | 0.03 | 0.02 | 0.04 | 0.00 | 0.03 | 0.01 | 0.003 | 0.38 (0.23-0.64) |
| Delayed hypersensitivity reaction | 0.40 | 0.27 | 0.52 | 0.04 | 0.43 | 0.36 | 0.044 | Not estimable<sup>e</sup> |
| Extensive limb swelling | 5.17 | 1.26 | 8.62 | 0.25 | 6.44 | 1.96 | 0.568 | Not estimable<sup>e</sup> |
| Facial swelling (in individuals with dermal fillers) | 4.99 | 1.76 | 7.69 | 0.62 | 4.04 | 4.03 | 0.548 | Not estimable<sup>e</sup> |
| Dizziness | 64.65 | 34.56 | 89.22 | 12.78 | 72.99 | 39.65 | 7.102 | Not estimable<sup>e</sup> |
| Tinnitus | 9.06 | 7.26 | 10.32 | 0.71 | 10.14 | 5.33 | 0.996 | 0.02 (0.02-0.02) |

CI, confidence interval; CLS, capillary leak syndrome; DIC, disseminated intravascular coagulation; OE, observed-to-expected.
<sup>a</sup>OE analyses were conducted using a 21-day risk window unless otherwise specified.
<sup>b</sup>Also includes target lesions and toxic skin eruption.
<sup>c</sup>Given limited feasibility of capturing subacute conditions in spontaneous adverse event reporting data, this outcome was defined as thyrotoxicosis.
<sup>d</sup>Systemic CLS is a rare disorder with a poorly characterized incidence rate. Fewer than 500 cases of CLS have been reported worldwide, and the prevalence of CLS has been reported as <1/1,000,000 [33, 50]. As such, validity of OE estimates is limited.
<sup>e</sup>For some outcomes for which population-based data characterizing a suitable expected event rate was not available, OE analyses could not be performed.

#### Myocarditis and/or pericarditis

For myocarditis, 4,723 cases were expected <21 days post-vaccination; 1,574 were expected <7 days. For pericarditis, 3,044 and 1,015 cases were expected <21 and <7 days, respectively. There were 6,702 cases (7,076 events) of myocarditis and pericarditis reported to the GSDB (0.7% and 0.5% of all cases, respectively). Most of the cases were reported in men aged 18-39 years (39.2%). Cases of myocarditis and pericarditis were most frequently reported after dose 2 (28.4%); the majority of cases had an onset <7 days after vaccination (65.5%). Most cases reported an outcome of recovered or recovering (46.9%), with 82 cases reporting a fatal outcome. Of those fatal cases, 5 included events of myopericarditis, 7 pericarditis, 66 myocarditis, 1 giant cell myocarditis, and 4 carditis. Most fatal cases occurred in men (66.3%), with a median age of 58 years and a median TTO of 5 days after any dose. Of the fatal cases, most had associated medical history (eg, cardiovascular diseases, COVID-19 infection) that may have contributed to the fatal outcome.

Myocarditis had an overall reporting rate of 6.07 cases per 1 million doses administered; pericarditis had an overall reporting rate of 4.72 cases per 1 million doses administered, with a higher rate in younger adults, as has been previously described (**Table 3**) [45]. For the overall population, the reporting rate of myocarditis was similar to the expected rate estimated in a US population-based study (OE ratio, 0.93; 95% CI, 0.90-0.97) when assuming a 21-day risk window. Myocarditis was reported at an increased rate within 7 days after vaccination (OE ratio, 1.41; 95% CI, 1.32-1.51), especially among individuals 12-40 years of age (3.51; 95% CI, 3.19-3.86) and after dose 2 among men 12-40 years of age ([post-dose 2] 9.75; 95% CI, 7.74-12.3). The rate of pericarditis was also elevated in individuals 12-40 years of age within 7 days after vaccination (2.54; 95% CI, 2.16-2.99).

#### TTS

For TTS, 845 cases were expected <21 days post-vaccination. There were 230 cases (250 events) identified with TTS-related PTs (0.02% of all cases); of those, 31 had a fatal outcome. No major differences were observed in cases reported in men (50.9%) compared with women (46.5%); the mean age was 59.5 years. When dose numbers and TTO were reported, cases were most frequently reported after dose 2 (29.2%) and >7 days after vaccination (96; 38.4%). Comparing the reporting rate to population-based estimates from the ACCESS project [43], the most conservative estimate of the OE ratio was 0.27 (95% CI, 0.24-0.31) based on an expected incidence (**Table 3**). No subgroup analyses for age or sex demonstrated an elevated reporting rate.

#### GBS

For GBS, 2,591 cases were expected <21 days post-vaccination. There were 683 cases (713 events) reported under GBS-related PTs (0.1% of total cases); of these, 673 were considered serious and 10 had fatal outcomes. There were more cases reported in men (51.1%) than women (46.7%), and most cases were reported in individuals aged >50 years. When dose numbers and TTO were reported, more cases occurred after dose 1 (24.3%) or dose 2 (23.7%) and >14 days after any dose (36.6%). The reporting rate for GBS was below a US population-based estimate of incidence (OE ratio, 0.26; 95% CI, 0.24-0.29; **Table 3**). No subgroup analyses for age or sex demonstrated an elevated reporting rate.

#### Other AESI

Among other AESI, reporting rates varied across subgroups defined by age and sex. Overall and in all subgroups considered, the estimated reporting rates for these conditions were below or compatible with the expected background incidence rate, and none met the prespecified threshold of a 95% CI lower bound >1 in this analysis. Medical review of AESI have not suggested any additional risks associated with the administration of mRNA-1273 (**Table 3**).

## Discussion

The COVID-19 pandemic has placed extraordinary demands on organizations charged with the public health response. By defining AESI in advance of authorization of COVID-19 vaccine use, SPEAC and the Brighton Collaboration provided a helpful framework to proactively anticipate and evaluate AESI that may occur in association with COVID-19 vaccines, whether incidentally or causally. To provide an update on the safety profile of mRNA-1273, we evaluated cumulative global AESI entered into the Moderna, Inc., GSDB from December 18, 2020, to December 17, 2022.

Given the large-scale use during the first 2 years after mRNA-1273 authorization (following the administration of >772 million doses), there was a very small percentage of fatalities (0.7%) following administration of mRNA-1273. Additionally, reports of certain AESI occurred at a relatively low rate, with 2,588 cases of anaphylaxis, 6,702 of myocarditis and/or pericarditis, 230 of TTS, and 683 of GBS. Our analysis confirmed an elevated risk for anaphylaxis, myocarditis, and/or pericarditis, but not for TTS or GBS. Upon confirmation of these risks, anaphylaxis, myocarditis, and pericarditis were added to mRNA-1273 prescribing information as part of good pharmacovigilance practices and to keep healthcare professionals and the general public informed.

An advance in pharmacovigilance made during the pandemic has been the use of background rates for conditions anticipated to be reported as AEs, allowing for a more intuitive approach to characterizing strength of association, and one more readily suited for additional analyses (eg, estimating strength of association categorized by age and sex). Previously, the quantitative evaluation of the strength of association for spontaneous reported data has relied upon disproportionality testing, a form of data mining that evaluates the strength of association of individual AEs observed following immunization compared with those observed following immunization with other vaccines [46]. In this analysis, OE ratios for most AESI evaluated did not exceed the threshold of the lower bound of the 95% CI >1 for the standard risk window of 21 days following vaccination. As has been previously reported [45, 47, 48], AESI of myocarditis with or without pericarditis and pericarditis demonstrated elevated reporting rates in subgroup analyses, with more marked associations observed in sensitivity analyses that restricted timing to a 7-day risk window after vaccination. Rates of anaphylaxis were also increased during the 3 days after vaccination.

Information presented in this analysis offers a safety evaluation of cases submitted to the GSDB. However, an inherent limitation is the spontaneous nature of cases received through the international voluntary reporting system. Moreover, when comparing observed reporting rates of AESI and expected background rates, there is a lack of visibility regarding patient-level data and limited availability of details on exposure in special risk groups who have received mRNA-1273. Additionally, published information on overall estimates of vaccine use varies, making it difficult to accurately estimate exposure to mRNA-1273, especially within population subgroups. Further, it should also be noted that many AESI have highly variable estimates of incidence across sources. Confounding almost certainly contributes to the variability as the proportion of mRNA-1273 vaccine recipients with relevant comorbidities and other risk factors for the outcomes assessed is unknown, and subgroup analyses of potential interest are infeasible. Some of these limitations may be mitigated by confirmation via observational studies evaluating specific safety questions. Nonetheless, post-marketing surveillance allows for the collection of information on a large number of individuals across a wide range of medical practices, provides data regarding unstudied uses or populations, and can detect rare, unexpected AEs [49]. There has been a notable reduction in cases submitted to the GSDB following the peak in May 2021, which likely reflects both an increasing familiarity with the safety profile of mRNA-1273, as well as the declining use of mRNA-1273 with the development and use of variant-containing vaccines.

## Conclusions

Among the list of AESI proposed by SPEAC in advance of COVID-19 vaccine use and with the exceptions of anaphylaxis, myocarditis, and pericarditis, this mRNA-1273 safety analysis did not find evidence to suggest an increased risk of AESI. Continuous safety monitoring of mRNA-1273 and variant-containing vaccines is ongoing.

## Acknowledgments

The authors would like to thank Oketoun Akangbe, Vasudev Bhupathi, Magalie Emilebacker, Kevin McNamara, Janaki Ram Neelakanti, and the rest of the pharmacovigilance scientists who supported the highlighted topics, along with Andrea Sutherland, Vaishali Khamamkar, and Samantha St Laurent, for their contributions to the manuscript. Medical writing and editorial assistance were provided by Jared Mackenzie, PhD, Ashlea Inan, PhD, and Lindsey Kirkland, PhD, of MEDiSTRAVA in accordance with Good Publication Practice guidelines (GPP 2022), funded by Moderna, Inc., and under the direction of the authors.

## Data Availability

Patient-level data reported in this study are not shared publicly but they are shared fully with regulatory agencies.

## Author Contributions

V.U., D.B.E., P.D., M.S.M., K.A., and W.S. conceived and designed the current study. V.U., D.B.E., P.D., and M.S.M. were involved with data collection. V.U., D.B.E., P.D., M.S.M., K.A., T.O.O., M.R., and W.S. carried out analysis and interpretation of data and take responsibility for the integrity of the data and accuracy of the data analysis. All authors critically reviewed and revised the paper for important intellectual content and approved the final draft of the manuscript.

## References

1. Centers for Disease Control and Prevention. COVID-19 Vaccines Work. Available at: https://www.cdc.gov/coronavirus/2019-ncov/vaccines/effectiveness/work.html#print.

2. World Health Organization. COVID-19 Advice for the Public: Getting vaccinated. Available at: https://www.who.int/emergencies/diseases/novel-coronavirus-2019/covid-19-vaccines/advice. Accessed 10 January 2023.

3. Scobie HM, Johnson AG, Suthar AB, et al. Monitoring Incidence of COVID-19 Cases, Hospitalizations, and Deaths, by Vaccination Status - 13 U.S. Jurisdictions, April 4-July 17, 2021. MMWR Morb Mortal Wkly Rep 2021; 70(37): 1284-90.

4. Johnson AG, Amin AB, Ali AR, et al. COVID-19 Incidence and Death Rates Among Unvaccinated and Fully Vaccinated Adults with and Without Booster Doses During Periods of Delta and Omicron Variant Emergence - 25 U.S. Jurisdictions, April 4-December 25, 2021. MMWR Morb Mortal Wkly Rep 2022; 71(4): 132-8.

5. Baden LR, El Sahly HM, Essink B, et al. Efficacy and safety of the mRNA-1273 SARS-CoV-2 vaccine. N Engl J Med 2021; 384(5): 403–16.

6. Chemaitelly H, Yassine HM, Benslimane FM, et al. mRNA-1273 COVID-19 vaccine effectiveness against the B.1.1.7 and B.1.351 variants and severe COVID-19 disease in Qatar. Nat Med 2021.

7. Bruxvoort KJ, Sy LS, Qian L, et al. Real-world effectiveness of the mRNA-1273 vaccine against COVID-19: Interim results from a prospective observational cohort study. Lancet Reg Health Am 2021: 100134.

8. Bruxvoort KJ, Sy LS, Qian L, et al. Effectiveness of mRNA-1273 against delta, mu, and other emerging variants of SARS-CoV-2: test negative case-control study. BMJ 2021; 375: e068848.

9. Embi PJ, Levy ME, Naleway AL, et al. Effectiveness of 2-dose vaccination with mRNA COVID-19 vaccines against COVID-19-associated hospitalizations among immunocompromised adults - nine states, January-September 2021. MMWR Morb Mortal Wkly Rep 2021; 70(44): 1553-9.

10. Bajema KL, Dahl RM, Evener SL, et al. Comparative Effectiveness and Antibody Responses to Moderna and Pfizer-BioNTech COVID-19 Vaccines among Hospitalized Veterans - Five Veterans Affairs Medical Centers, United States, February 1-September 30, 2021. MMWR Morb Mortal Wkly Rep 2021; 70(49): 1700-5.

11. World Health Organization. The Moderna COVID-19 (mRNA-1273) vaccine: what you need to know. Available at: https://www.who.int/news-room/feature-stories/detail/the-moderna-covid-19-mrna-1273-vaccine-what-you-need-to-know. Accessed March 14.

13. Our World in Data. COVID-19 Vaccine Doses Administered by Manufacturer. Available at: https://ourworldindata.org/grapher/covid-vaccine-doses-by-manufacturer. Accessed 7 March 2023.

13. Klein NP, Lewis N, Goddard K, et al. Surveillance for Adverse Events After COVID-19 mRNA Vaccination. JAMA 2021; 326(14): 1390–9.

14. World Health Organization. COVID-19 Vaccines: Safety Surveillance Manual, 2020.

15. Shimabukuro TT. COVID-19 vaccine post-authorization safety monitoring update, 2020.

16. VAC4EU. COVID-19 vaccine monitoring. Available at: https://vac4eu.org/covid-19-vaccine-monitoring/. Accessed April 3.

17. Younus MM, Al-Jumaili AA. An Overview of COVID-19 Vaccine Safety and Post-marketing Surveillance Systems. Innov Pharm 2021; 12(4).

19. Safety Platform for Emergency vACcines (SPEAC). Priority List of Adverse Events of Special Interest: COVID-19. Available at: https://media.tghn.org/articles/COVID-19_AESIs_SPEAC_V1.1_5Mar2020.pdf. Accessed April 3.

19. Oster ME, Shay DK, Su JR, et al. Myocarditis Cases Reported After mRNA-Based COVID-19 Vaccination in the US From December 2020 to August 2021. JAMA 2022; 327(4): 331–40.

20. Centers for Disease Control and Prevention. Safety of COVID-19 Vaccines. Available at: https://www.cdc.gov/coronavirus/2019-ncov/vaccines/safety/safety-of-vaccines.html. Accessed March 14.

21. CDC COVID-19 Response Team; Food and Drug Administration. Allergic Reactions Including Anaphylaxis After Receipt of the First Dose of Moderna COVID-19 Vaccine - United States, December 21, 2020-January 10, 2021. MMWR Morb Mortal Wkly Rep 2021; 70(4): 125-9.

22. Shimabukuro T, Nair N. Allergic Reactions Including Anaphylaxis After Receipt of the First Dose of Pfizer-BioNTech COVID-19 Vaccine. JAMA 2021; 325(8): 780–1.

23. Long B, Bridwell R, Gottlieb M. Thrombosis with thrombocytopenia syndrome associated with COVID-19 vaccines. Am J Emerg Med 2021; 49: 58–61.

24. Hanson KE, Goddard K, Lewis N, et al. Incidence of Guillain-Barre Syndrome After COVID-19 Vaccination in the Vaccine Safety Datalink. JAMA Netw Open 2022; 5(4): e228879.

25. Mahaux O, Bauchau V, Van Holle L. Pharmacoepidemiological considerations in observed-to-expected analyses for vaccines. Pharmacoepidemiol Drug Saf 2016; 25(2): 215–22.

26. Safety Platform for Emergency vACcines (SPEAC). Updated (October 2022) COVID-19 AESI including status of associated Brighton case definitions, 2022.

27. International Council for Harmonisation of Technical Requirements for Pharmaceuticals for Human Use (ICH). Clinical Safety Data Management: Definitions and Standards for Expedited Reporting E2A, 1994.

28. United States Food and Drug Administration. CFR - Code of Federal Regulations Title 21. Available at: https://www.accessdata.fda.gov/scripts/cdrh/cfdocs/cfcfr/cfrsearch.cfm?fr=312.32. Accessed June 15, 2023.

29. Centers for Disease Control and Prevention. COVID-19 Vaccinations in the United States. Available at: https://covid.cdc.gov/covid-data-tracker/#vaccinations_vacc-total-admin-rate-total. Accessed April 29, 2022.

30. Arora A, Wetter DA, Gonzalez-Santiago TM, Davis MD, Lohse CM. Incidence of leukocytoclastic vasculitis, 1996 to 2010: a population-based study in Olmsted County, Minnesota. Mayo Clin Proc 2014; 89(11): 1515-24.

31. Boehmer TK, Kompaniyets L, Lavery AM, et al. Association between COVID-19 and myocarditis using hospital-based administrative data - United States, March 2020-January 2021. MMWR Morb Mortal Wkly Rep 2021; 70(35): 1228-32.

32. Chen J, Tian DC, Zhang C, et al. Incidence, mortality, and economic burden of myasthenia gravis in China: A nationwide population-based study. Lancet Reg Health West Pac 2020; 5: 100063.

33. Druey KM, Parikh SM. Idiopathic systemic capillary leak syndrome (Clarkson disease). J Allergy Clin Immunol 2017; 140(3): 663–70.

34. Esposito D, Titievsky L, Beachler DC, et al. Incidence of outcomes relevant to vaccine safety monitoring in a US commercially-insured population. Vaccine 2018; 36(52): 8084–93.

35. Kumar N, Pandey A, Jain P, Garg N. Acute Pericarditis-Associated Hospitalization in the USA: A Nationwide Analysis, 2003-2012. Cardiology 2016; 135(1): 27-35.

36. Li X, Ostropolets A, Makadia R, et al. Characterising the background incidence rates of adverse events of special interest for covid-19 vaccines in eight countries: multinational network cohort study. BMJ 2021; 373: n1435.

37. Marrie RA, Yu BN, Leung S, et al. The incidence and prevalence of fibromyalgia are higher in multiple sclerosis than the general population: A population-based study. Mult Scler Relat Disord 2012; 1(4): 162–7.

38. Martinez C, Wallenhorst C, McFerran D, Hall DA. Incidence rates of clinically significant tinnitus: 10-year trend from a cohort study in England. Ear Hear 2015; 36(3): e69–75.

39. Mount HR, Boyle SD. Aseptic and Bacterial Meningitis: Evaluation, Treatment, and Prevention. Am Fam Physician 2017; 96(5): 314–22.

40. National Organization for Rare Disorders (NORD). Systemic Capillary Leak Syndrome. Available at: https://rarediseases.org/rare-diseases/systemic-capillary-leak-syndrome/. Accessed May 18.

41. Otite FO, Patel S, Sharma R, et al. Trends in incidence and epidemiologic characteristics of cerebral venous thrombosis in the United States. Neurology 2020; 95(16): e2200–e13.

42. Weycker D, Hanau A, Hatfield M, et al. Primary immune thrombocytopenia in US clinical practice: incidence and healthcare burden in first 12 months following diagnosis. J Med Econ 2020; 23(2): 184–92.

43. Willame C, Dodd C, Duran CE, et al. Background rates of 41 adverse events of special interest for COVID-19 vaccines in 10 European healthcare databases - an ACCESS cohort study. Vaccine 2023; 41(1): 251–62.

44. Yadav D, Timmons L, Benson JT, Dierkhising RA, Chari ST. Incidence, prevalence, and survival of chronic pancreatitis: a population-based study. Am J Gastroenterol 2011; 106(12): 2192–9.

45. Straus W, Urdaneta V, Esposito DB, et al. Analysis of Myocarditis Among 252 Million mRNA-1273 Recipients Worldwide. Clin Infect Dis 2023; 76(3): e544–e52.

46. Montastruc JL, Sommet A, Bagheri H, Lapeyre-Mestre M. Benefits and strengths of the disproportionality analysis for identification of adverse drug reactions in a pharmacovigilance database. Br J Clin Pharmacol 2011; 72(6): 905–8.

47. Wong HL, Hu M, Zhou CK, et al. Risk of myocarditis and pericarditis after the COVID-19 mRNA vaccination in the USA: a cohort study in claims databases. Lancet 2022; 399(10342): 2191–9.

48. Gao J, Feng L, Li Y, et al. A Systematic Review and Meta-analysis of the Association Between SARS-CoV-2 Vaccination and Myocarditis or Pericarditis. Am J Prev Med 2023; 64(2): 275–84.

49. Kennedy DL, Goldman SA, Lillie RB. Spontaneous Reportin in the United States. In: Strom BL. Pharmacoepidemiology Vol. 3rd ed. Chicester, United Kingdom: Wiley, 2000:151-74.

50. Kapoor P, Greipp PT, Schaefer EW, et al. Idiopathic systemic capillary leak syndrome (Clarkson’s disease): the Mayo clinic experience. Mayo Clin Proc 2010; 85(10): 905–12.

